# The Effectiveness of Transcranial Magnetic Stimulation in Suicidality: An Updated Systematic Review

**DOI:** 10.1101/2025.05.05.25327034

**Authors:** S. J. Manuele, A. Cerins, L. M. Jenkins, E. H. X. Thomas, N. Sabetfakhri, E. Cholakians, A. L. Aaronson, L. Chen

## Abstract

Suicidal thoughts and behaviors (STB) have a substantial global burden, with over 14 million individuals attempting suicide annually. Existing biological therapies do not adequately reduce STB risk. Repetitive transcranial magnetic stimulation (rTMS) is an approved, non-invasive and low-risk treatment for several psychiatric disorders.

Meta-analyses investigating TMS’ effects on STB indicate therapeutic promise. Given the proliferation of TMS studies investigating its effect on STB, a repeat review of the literature is warranted.

**Methods:** A PRISMA-guided systematic review was conducted to evaluate the efficacy of rTMS in reducing STB. Studies assessing STB outcomes following rTMS to treat psychiatric disorders, either as monotherapy or adjunctive treatment, were included. Forty-five studies were identified (*N*=3515).

**Results:** Studies generally applied rTMS to treat primary psychiatric disorders, particularly depression, with change in suicidality evaluated as a secondary outcome. rTMS protocols differed across studies. Most studies targeted the left dorsolateral prefrontal cortex (dlPFC), although significant improvements to STB were also reported with rTMS targeting the visual cortex, right dlPFC and the bilateral PFC. High frequency rTMS and intermittent theta burst stimulation (iTBS) protocols were superior in reducing STB compared to other stimulation protocols, with studies reporting 40-100% treatment response rates. Adverse events (AEs) were mostly mild and transient.

**Conclusion:** rTMS appears to be a safe and effective treatment option for STB, with significant reductions observed, particularly when rTMS or iTBS is applied to the dlPFC. Mechanistically informed randomized controlled trials specifically designed to evaluate rTMS’ treatment effects on STB are needed to validate this promising treatment approach.

## Introduction

Globally, more than 14.5 million people attempt suicide each year, with over 725,000 deaths resulting from suicide (*World Health Organization*, 2024). Suicidal thoughts and behaviors (STB) can occur in the context of nearly any psychiatric diagnosis, but can also occur in individuals without a psychiatric disorder (Caudle et al., 2024). STB have been linked to a range of risk factors, including stress, maladaptive emotion and pain regulation, reward seeking, and cognitive and inhibitory control deficits that often occur from a range of mental and physical health problems, as well as environmental stressors (Abdollahpour Ranjbar et al., 2024; Barredo et al., 2021).

There are biological, psychological and behavioral strategies to manage STB though the efficacy of each strategy is limited. Pharmacotherapy (e.g., antidepressants, mood stabilisers, antipsychotic medications and ketamine compounds) is often used (Zisook et al., 2023). However, while Ketamine studies have shown promise in rapidly alleviating STB, side effects varied (e.g., sedation, depersonalization, agitation, nausea) and benefits were reported to last up to two weeks (Abbar et al., 2022; Bruton et al., 2025; Wilkinson et al., 2018). The short window of symptom alleviation may result in post-ketamine treatment STB spikes (Ingrosso et al., 2025; Lascelles et al., 2021). Further, A recent meta-analysis examining the effects of the ketamine enantiomer, esketamine, on STB reported limited efficacy. Electroconvulsive Therapy (ECT), an evidence-based and relatively rapid-acting treatment for several psychiatric conditions, may be used as an STB treatment (Salik & Marwaha, 2025). However, stigma and concerns regarding cognitive and memory side effects may limit its acceptance as a treatment option (Argyelan et al., 2021; Porter et al., 2020). Further, ECT requires the administration of general anaesthesia, thus exposing those receiving ECT to its inherent risks (Salik & Marwaha, 2025). Implications to blood pressure, heart rate and intracranial pressure may contraindicate ECT for those with central nervous system, respiratory and cardiovascular issues (Salik & Marwaha, 2025). Evidence for the effectiveness of manualized psychological therapies in treating STB, namely cognitive-behavioral therapy and dialectical behavior therapy, appears mixed (Sufrate-Sorzano et al., 2023).

Overall, meta-analyses suggest existing STB treatments have varying efficacy.

One study examined 29 placebo-controlled trials, ultimately concluding there was inadequate evidence to support antidepressants, as a treatment to prevent STB (Braun et al., 2016). Antidepressant interventions for STB are not immediate acting, creating vulnerability for those with active STB (Del Matto et al., 2020).

Pharmacotherapy often requires trialling of numerous medications to experience symptom reduction, while remission is often dosage dependent. Ongoing continuation or maintenance, as well as strict adherence to guidelines for ECT, ketamine and pharmacotherapy are often necessary but underutilized for sustained benefit (Jørgensen et al., 2024). The limitations of current interventions highlight a critical gap in effective, tolerable treatments for suicidality – one that emerging neuromodulation techniques may begin to address. Thus, there is a critical unmet need for innovative treatments that are both effective and tolerable for reducing suicidality.

Repetitive transcranial magnetic stimulation (rTMS), a non-invasive brain stimulation technique, is approved by the Food and Drug Administration (U.S. FDA) as an evidence-based treatment for several neuropsychiatric conditions, such as major depression (Cohen et al., 2022; Mutz et al., 2018). These studies target the dorsolateral prefrontal cortex (dlPFC) due to its implication in mood symptoms.

Emerging literature report the effects of stimulation applied to target the motor, anterior cingulate and orbitofrontal cortices as treatment sites for other psychiatric conditions, many of which may have heightened STB risk (Han et al., 2023; Lusicic et al., 2018).

There is a pressing need for novel treatment strategies that can reduce STB. An expanding body of clinical trial data suggests that rTMS may effectively reduce STB. The most recent review of the literature reporting on rTMS’s efficacy in treating STB occurring transdiagnostically was published in 2022 (G.-W. Chen et al., 2022). A substantial number of studies of this promising approach to alleviating suicidal ideation have been published since (Aaronson et al., 2024; Adu et al., 2023; Hickson et al., 2024; Huang et al., 2025; Kong et al., 2023; Li et al., 2024; Pan et al., 2023; Sun et al., 2024; Terpstra et al., 2023; Thai et al., 2024; Wilkinson et al., 2023; Zhan et al., 2024; Zhao et al., 2024; Zhao et al., 2023). Given the volume of new data available and the urgent need for novel intervention strategies, this systematic review provides an updated insight into the current literature investigating the efficacy of rTMS in individuals experiencing STB. Further, this review aims to address the efficacy of various rTMS modalities and protocols on STB symptoms including suicidal ideation, attempts, and behaviors.

## Methods

A systematic review of the existing literature regarding the efficacy of TMS in individuals with STB was conducted, utilizing the Preferred Reporting Items for Systematic Reviews and Meta-Analyses (PRISMA) guidelines (Page, McKenzie, et al., 2021; Page, Moher, et al., 2021).

### Search Strategy

Using PubMed and Web of Science databases, a literature search was conducted. Search terms included suicid*, transcranial magnetic stimulation, TMS.

To ensure rigorous assessment of the existing literature, publications were not limited by date. The final study search was completed on February 12, 2025. Studies which were published until February 2025 were included. See Figure 1 for PRISMA flow diagram.

**Figure 1.**
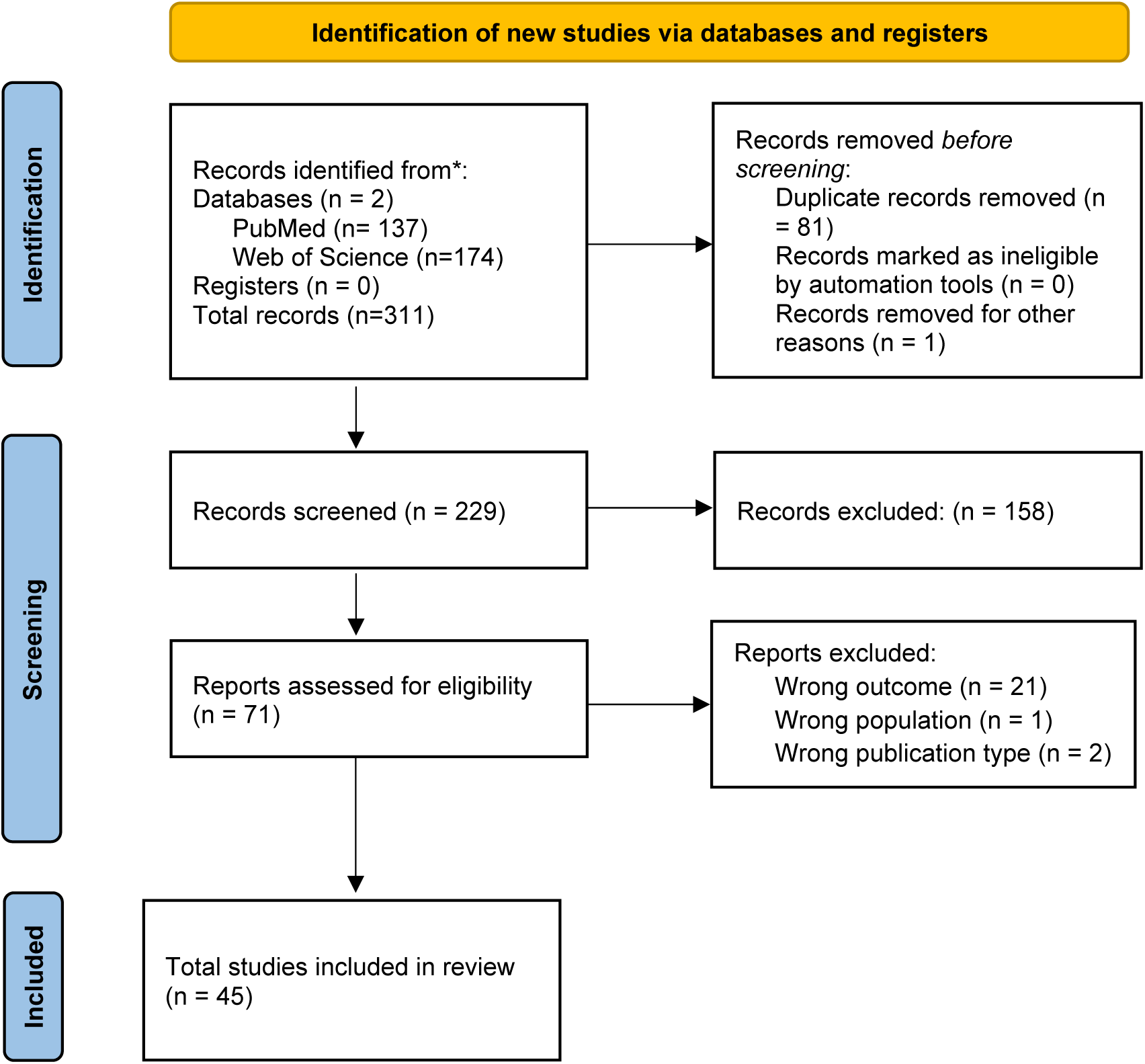
PRISMA Flow Diagram

### Inclusion Criteria and Study Selection

*Intervention:* Studies which assessed TMS independently or as an adjunct treatment for STB were included. This review included studies regardless of TMS site, type (i.e. Repetitive [rTMS]; Deep [dTMS], Intermittent Theta Burst [iTBS], Accelerated iTBS, Low Frequency rTMS [LFR rTMS] or Continuous Theta Burst [cTBS]), trial duration, frequency (Hz) or number of pulses delivered.

*Outcome Measures:* Studies which assessed STB as a primary or secondary outcome were included if measures of STB were collected at multiple timepoints. In addition, studies which measured self-or clinician-rated, or in the instance of youth studies, parent-rated STB, were included.

*Population:* Studies which included participants who reported STB. Studies were not excluded if they had participants with comorbid psychiatric conditions. No exclusion criteria were specified for age of participants, or concurrent or past treatment or intervention methods.

*Comparison/Placebo/Control Group:* Studies considered for this review were not required to have recruited a comparison or control group or have administered sham TMS.

*Study Type:* Studies which used observational or interventional designs, such as randomized controlled trials, clinical trials, cohort studies, open-label studies, pilot studies and case-control studies, were included. Individual case reports, reviews and summary publications were excluded. Studies were also required to be published and/or available in English. Gray literature and published dissertations were excluded.

### Data Screening

Rayyan software (https://www.rayyan.ai/; Ouzzani et al., 2016) was utilized to screen the identified literature. After duplicate screening and exclusion by Rayyan, study screening was conducted by AC and SJM, independently. Full text retrieval was conducted for manuscripts which met inclusion criteria, and full text screening was completed by SJM. Disagreements were resolved by AC and SJM. Studies excluded at full text screening stage and exclusion reason are listed in Figure 1.

### Data Extraction

Information from included studies was extracted, including study design, intervention details such as protocol, frequency, location and number of TMS treatments, use of neuronavigation for treatment, adverse events or side effects of treatment, demographic information, baseline and follow up symptomatology, and pre-existing comorbidities. See Supplementary Materials for data extraction table.

## Results

A database search yielded 311 studies for consideration, of which 81 were identified as duplicates. The remaining 229 articles were title and abstract screened. We excluded 158 articles at the title and abstract screen phase, resulting in 71 full texts screened. The final number of studies included in the review was 45, with most studies excluded at the full text screening stage due to incorrect outcome measured (namely depressive symptoms which encompassed STB but did not address change in STB independently). A PRISMA flow chart is presented in Figure 1. The 45 included studies encompassed a total sample size of 3,515 participants, ranging in age from 13 to 80 years. Most studies however, included adult samples, with mean ages between 40 to 55. Additionally, of the 44 studies (*n*=3483) that provided sex distribution of the samples, 67.4% of participants were female.

The studies included in this review implemented various TMS modalities, including repetitive TMS (rTMS), deep TMS (dTMS), intermittent theta burst stimulation (iTBS), continuous theta burst stimulation (cTBS), and accelerated protocols for iTBS and cTBS. Participants enrolled in these studies had existing diagnoses of TRD (*n*=22), Depression or MDD (*n*=20), bipolar disorder (BD) (*n*=2), BPD (*n*=1), PTSD (*n*=3), and/or Traumatic Brain Injury (TBI) (*n*=2). While most studies treated the left dlPFC, other targets included the visual cortex (VC), right dlPFC, dorsomedial prefrontal cortex (dmPFC), and the bilateral dlPFC. Additionally, the anterior cingulate cortex (ACC) was targeted with dTMS. STB measures varied substantially across studies, with some administering STB specific questionnaires (e.g., Columbia Suicidal Severity Rating Scale [C-SSRS]; Beck Suicidality Index [BSI]), and others extracting suicide-related items from broader questionnaires (e.g., Hamilton Depression Rating Scale Item 3, [HAMD]; Montgomery-Asberg Depression Rating Scale Item 10 [MADRS]). Finally, protocols for identifying TMS treatment site varied, with studies using neuronavigation (*n*=17), participant anatomical scalp measures (*n*=15), BeamF3 (*n*=2), and some unspecified, while H1 coils were implemented for dTMS protocols (*n*=2). Scalp measured varied in distance from reference region, and reference region. See Table 1 for study details.

**Table 1.**
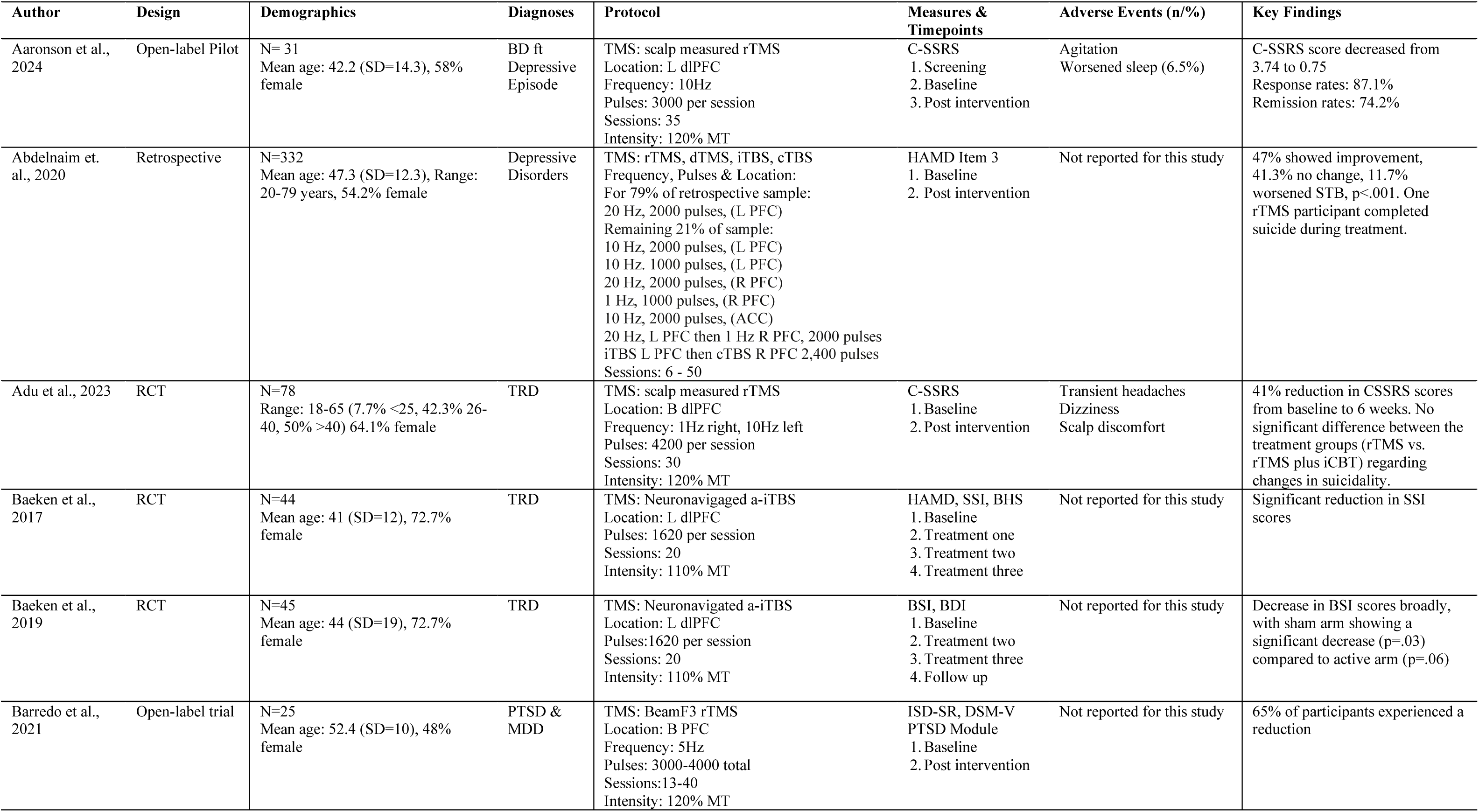
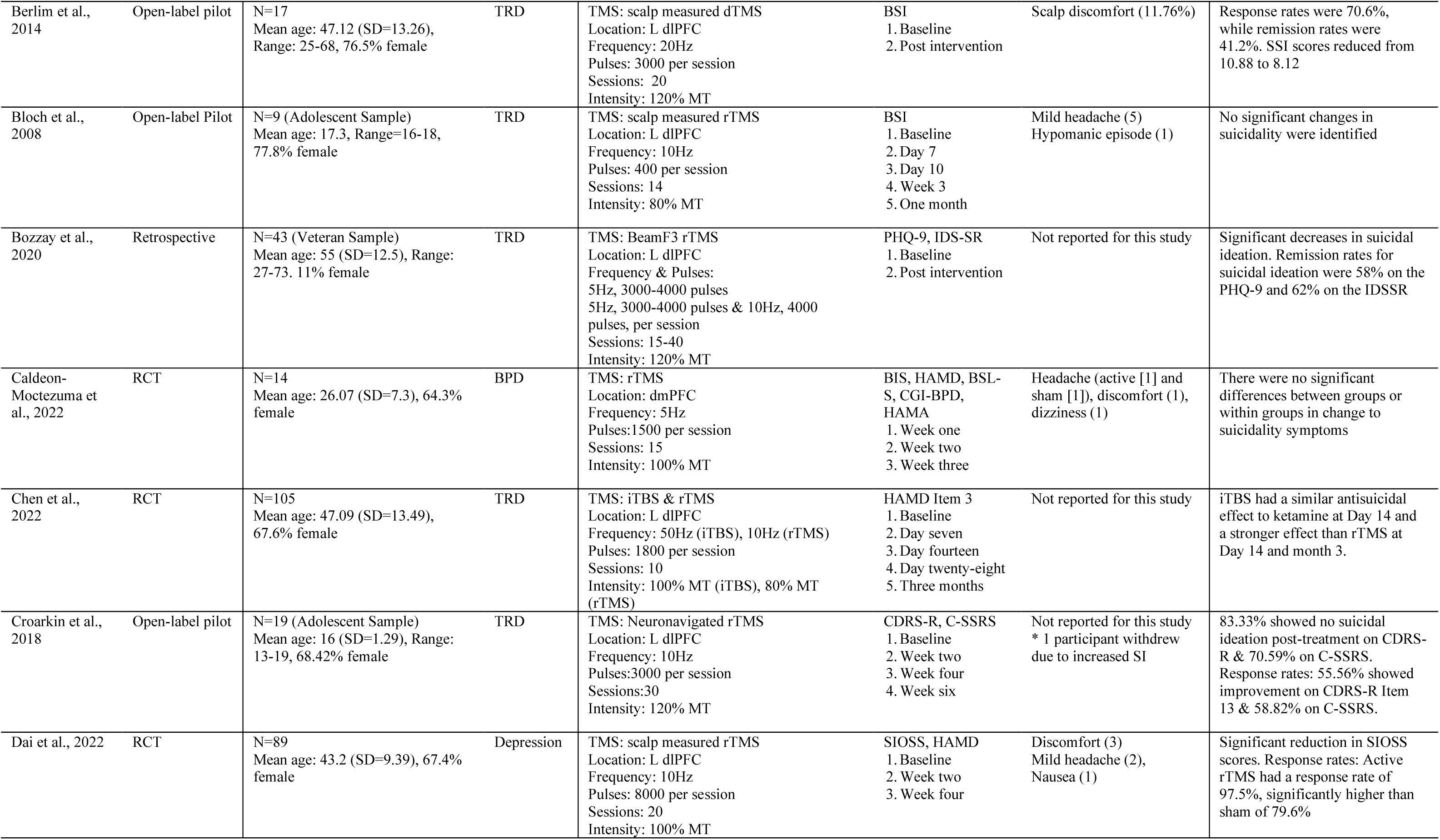
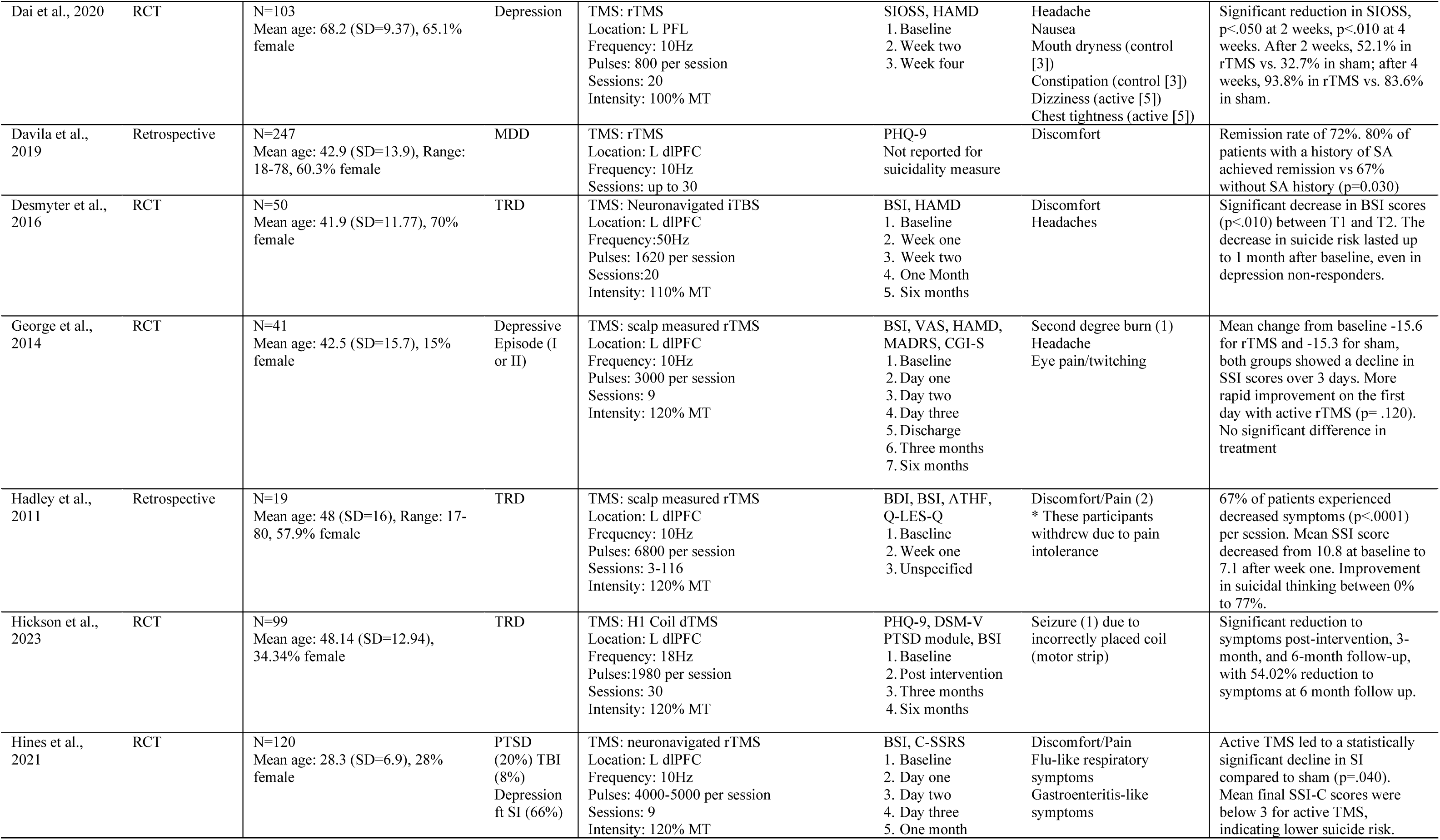
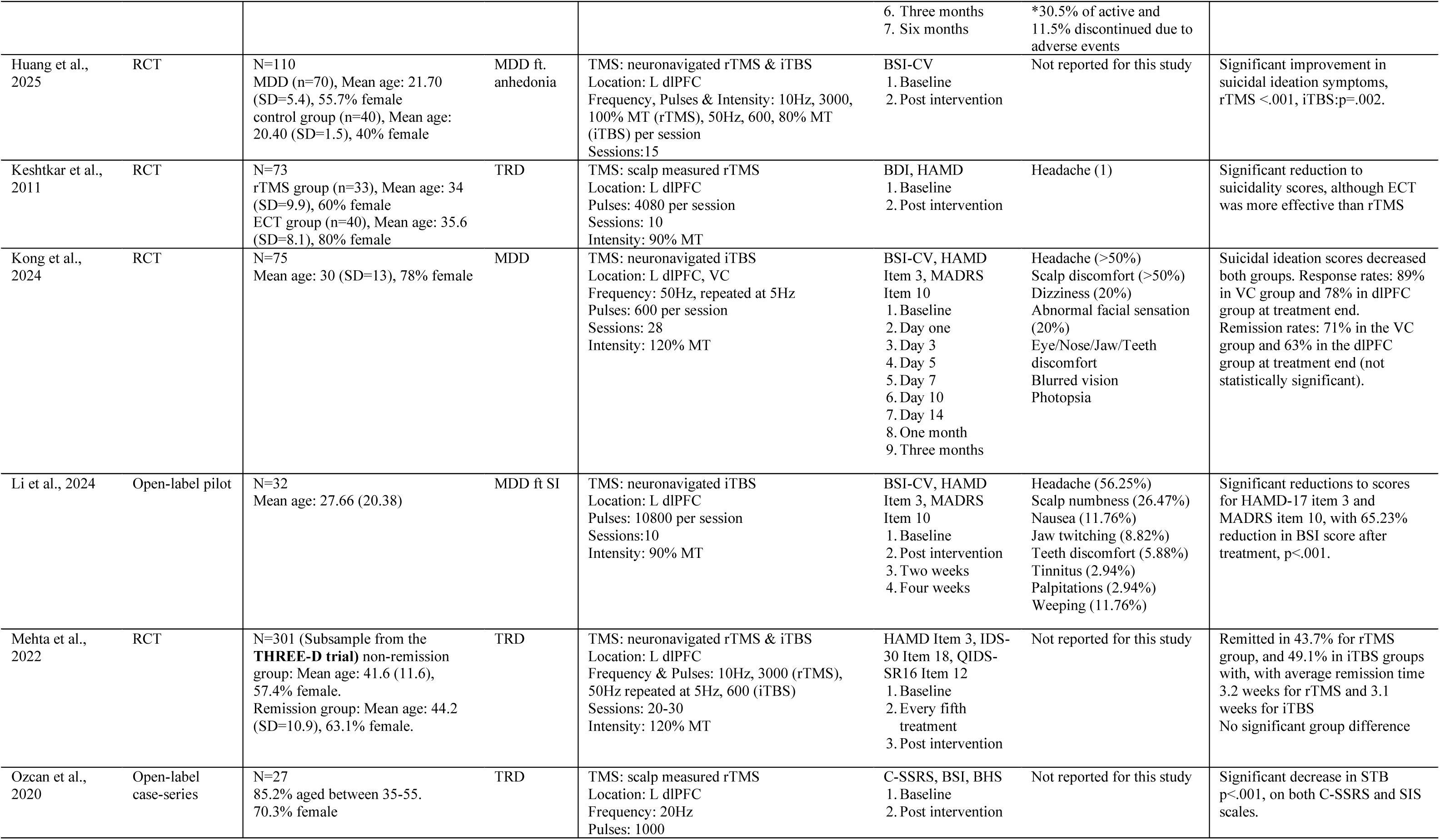
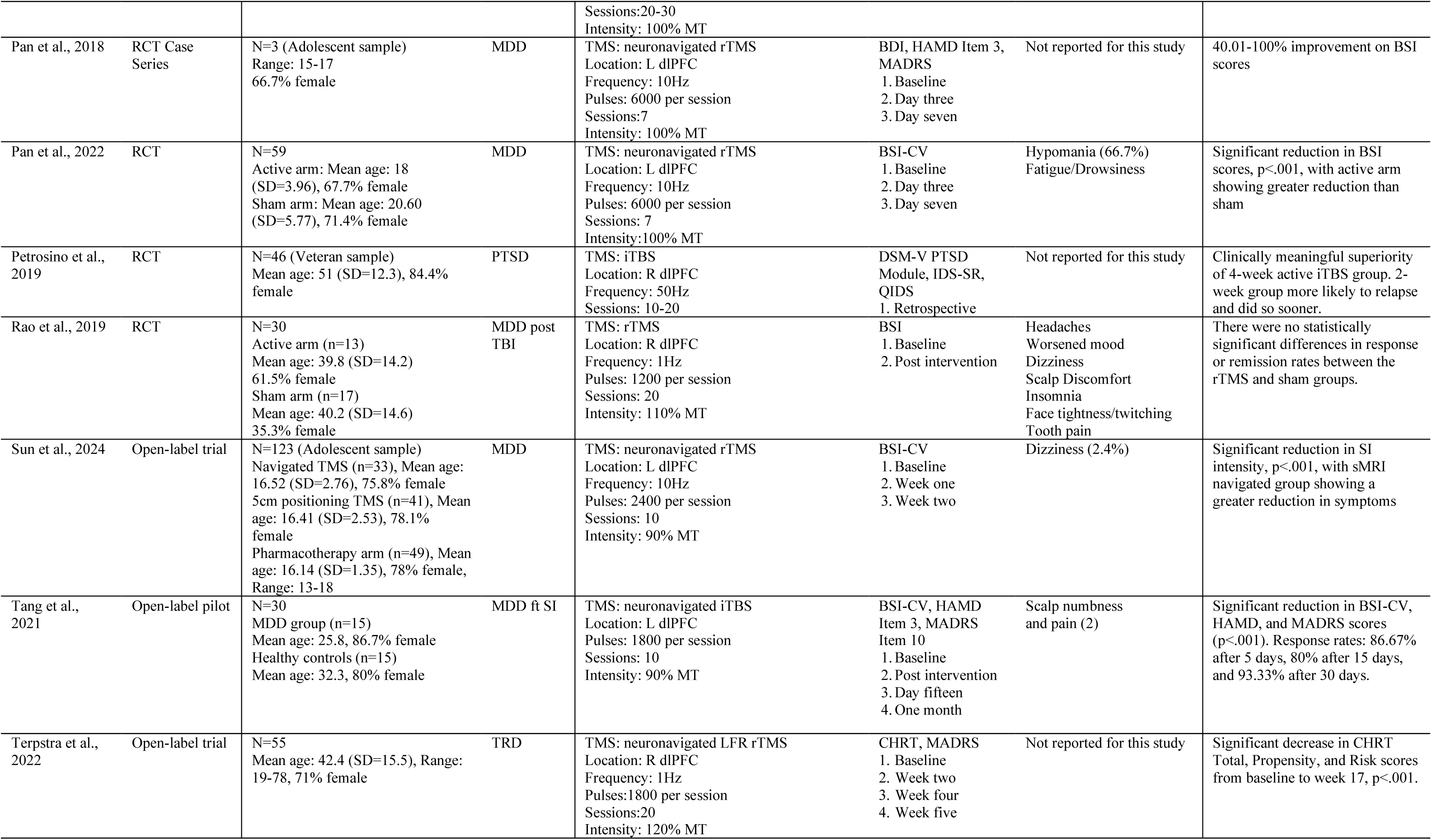
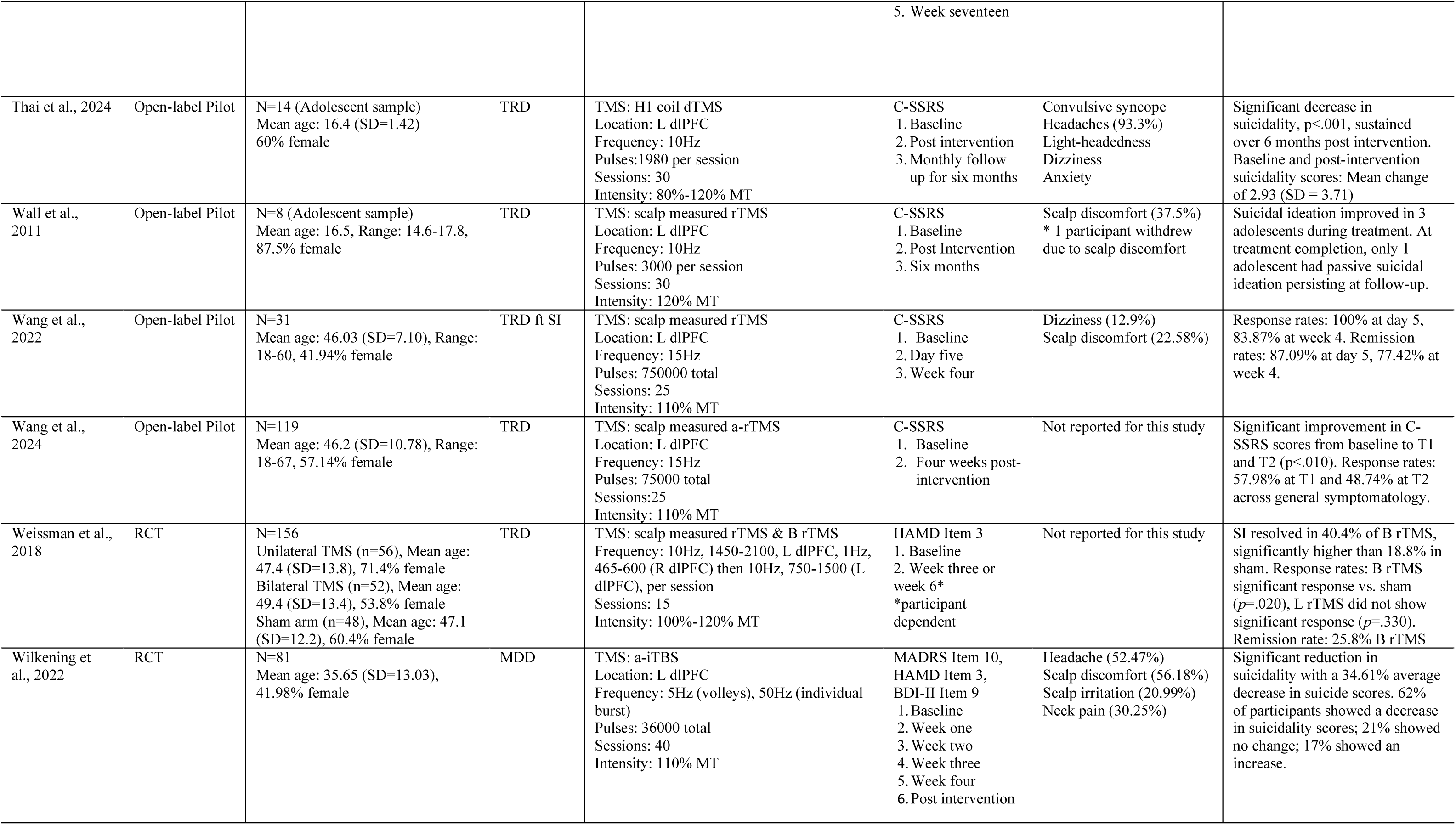
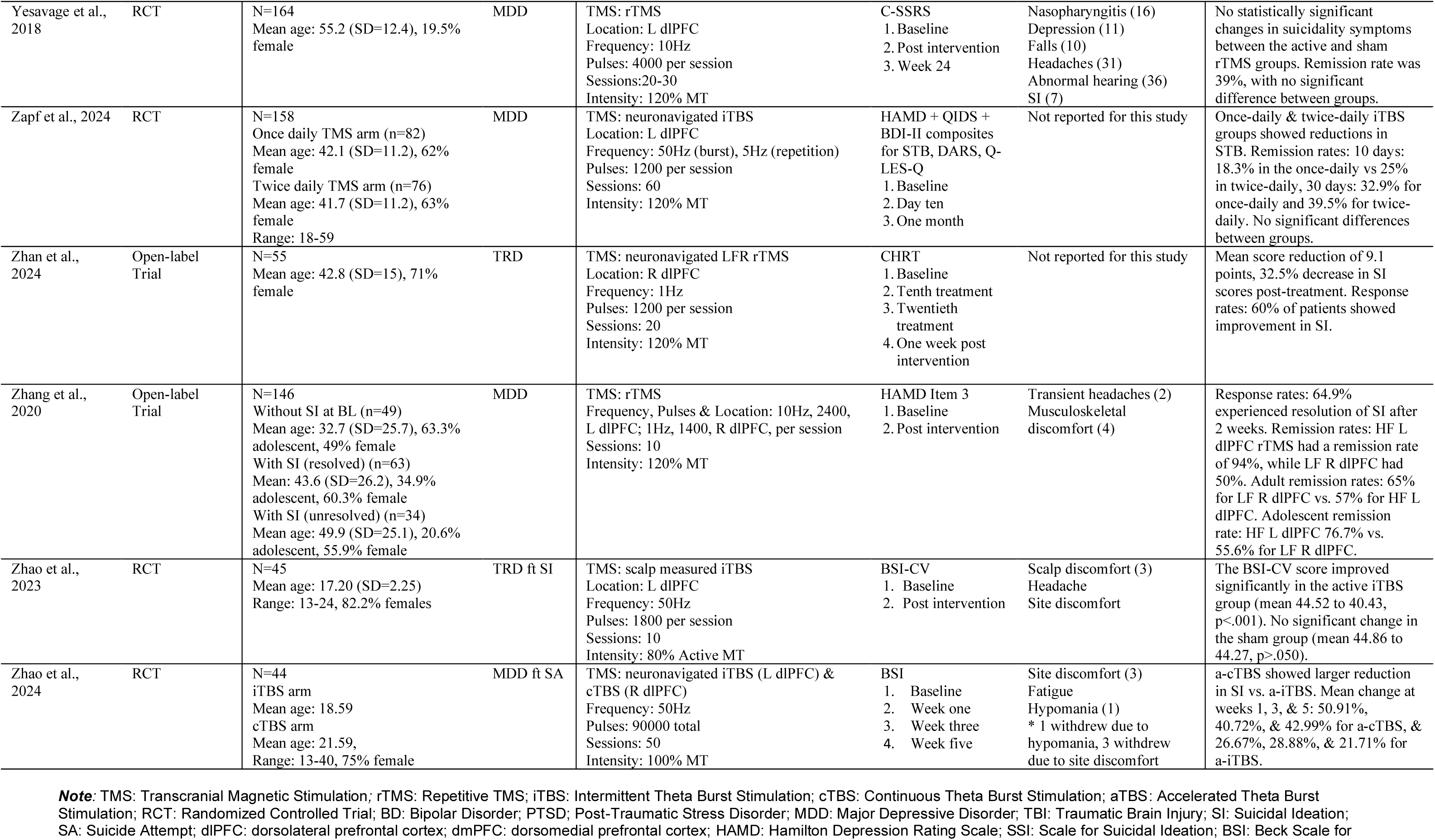
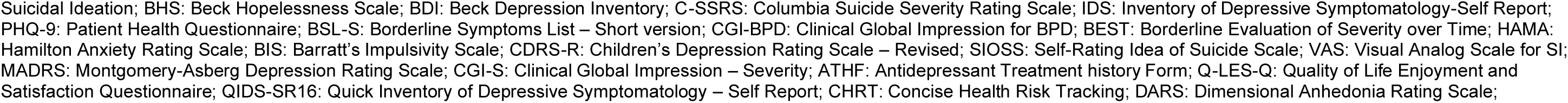
Study details.

### TMS to the prefrontal cortex (PFC)

Forty-one of the identified studies targeted the PFC with TMS for STB. See Table 2 for an overview of TMS protocols and outcomes for STB.

**Table 2.**
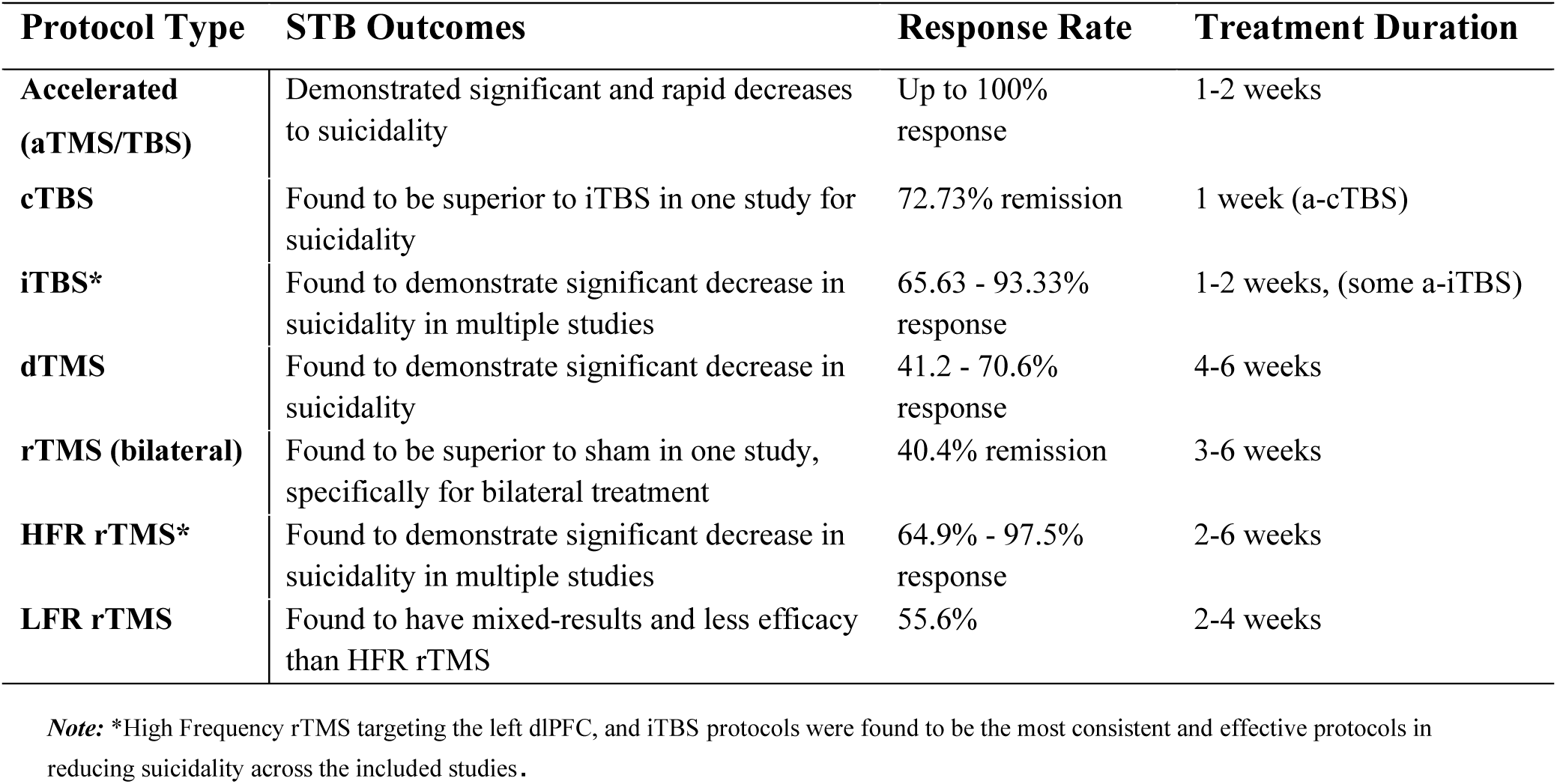
TMS Protocols and Outcomes for STB.

| Protocol Type | STB Outcomes | Response Rate | Treatment Duration |
| --- | --- | --- | --- |
| **Accelerated (aTMS/TBS)** | Demonstrated significant and rapid decreases to suicidality | Up to 100% response | 1-2 weeks |
| **cTBS** | Found to be superior to iTBS in one study for suicidality | 72.73% remission | 1 week (a-cTBS) |
| **iTBS\*** | Found to demonstrate significant decrease in suicidality in multiple studies | 65.63 - 93.33% response | 1-2 weeks, (some a-iTBS) |
| **dTMS** | Found to demonstrate significant decrease in suicidality | 41.2 - 70.6% response | 4-6 weeks |
| **rTMS (bilateral)** | Found to be superior to sham in one study, specifically for bilateral treatment | 40.4% remission | 3-6 weeks |
| **HFR rTMS\*** | Found to demonstrate significant decrease in suicidality in multiple studies | 64.9% - 97.5% response | 2-6 weeks |
| **LFR rTMS** | Found to have mixed-results and less efficacy than HFR rTMS | 55.6% | 2-4 weeks |

*Note:* \*High Frequency rTMS targeting the left dlPFC, and iTBS protocols were found to be the most consistent and effective protocols in reducing suicidality across the included studies.

### rTMS applied to the left dlPFC for reducing STB

Of the 41 studies which targeted the PFC, 36 targeted the left dlPFC. Twenty-three of these implemented rTMS protocols, with frequencies between 5Hz-20Hz (Aaronson et al., 2024; Abdelnaim et al., 2019; Bloch et al., 2008; Bozzay et al., 2020; M. H. Chen et al., 2022; Croarkin et al., 2018; Dai et al., 2022; Davila et al., 2019; George et al., 2014; Hadley et al., 2011; Hines et al., 2022; Huang et al., 2025; Keshtkar et al., 2011; Mehta et al., 2022; Ozcan et al., 2020; Pan et al., 2018; Pan et al., 2023; Sun et al., 2024; Wall et al., 2011; Wang et al., 2022; Wang et al., 2024; Weissman et al., 2018; Yesavage et al., 2018; Zhang et al., 2021). Left dlPFC rTMS reduced STB symptoms, with improvements noted in all but two studies (Bloch et al., 2008; Weissman et al., 2018). Specifically, rTMS response rates for STB were indicated between 40-100% (Aaronson et al., 2024; Croarkin et al., 2018; Dai et al., 2022; Hadley et al., 2011; Wang et al., 2022; Wang et al., 2024; Zhang et al., 2021). Further, studies indicated remission rates between 39-94% (Aaronson et al., 2024; Bozzay et al., 2020; Croarkin et al., 2018; Davilla et al., 2019; Mehta et al., 2022; Wang et al., 2018; Yesevage et al., 2018; Zhang et al., 2020).

Additionally, of the 36 studies which targeted the left dlPFC, 13 studies implemented TBS protocols (iTBS, cTBS and aTBS), all of which used 50Hz frequency, some with 5Hz repetitions (Baeken et al., 2017; Baeken et al., 2019; M. H. Chen et al., 2022; Desmyter et al., 2016; Huang et al., 2025; Kong et al., 2023; Li et al., 2024; Mehta et al., 2022; Tang et al., 2021; Wilkening et al., 2022; Zapf et al., 2024; Zhao et al., 2024; Zhao et al., 2023). All TBS studies indicated improvements to STB. However, (Baeken et al., 2017; Baeken et al., 2019) noted that while there were improvements to STB across active and sham arms, the iTBS group (*p*=.06) did not show significant improvements compared to sham (*p*=.03). Of the TBS studies that provided response rates, 62-93.33% responded to treatment (Kong et al., 2023; Tang et al., 2021; Wilkening et al., 2022). Some TBS studies also indicated remission in 18.3-63% of participants (Kong et al., 2023; Mehta et al., 2022; Zapf et al., 2024). However, one study also indicated that STB worsened in 17% of participants (Wilkening et al., 2022).

Three studies reported the use of dTMS applied to the left dlPFC at a frequency of 10-20Hz (Berlim et al., 2014; Hickson et al., 2024; Thai et al., 2024). All studies indicated improvements to STB symptoms during and at completion of treatment, with Berlim and colleagues (2014) also indicating response in 70.6% of participants, and remission in 41.2% of participants.

### Right dlPFC

Six studies targeted the right dlPFC, four of which used low frequency (LF) 1Hz rTMS (Rao et al., 2019; Terpstra et al., 2023; Zhan et al., 2024; Zhang et al., 2021) and the remaining used TBS at a frequency of 50Hz (Petrosino et al., 2020; Zhao et al., 2024). The LF rTMS studies identified significant improvements in STB, however Rao and colleagues (2019) did not find significant group differences between the active arm and the sham arm. One study indicated a LF rTMS response rate of 60% (Zhan et al., 2024) while remission rates were 50-55.6% in another (Zhang et al., 2021).

The right dlPFC TBS studies both identified improvements in STB. Further, Zhao et al. (2024) used cTBS, finding it to be superior in reducing STB than left dlPFC iTBS. Futher, Petrosino et al. (2020) found that iTBS treatment for four weeks was superior to treatment for two weeks, with the two-week iTBS group more likely to have STB relapse and do so sooner than the four-week group.

### Bilateral dlPFC

Two studies used rTMS to bilaterally target the dlPFC (Adu et al., 2023; Weissman et al., 2018). Both studies applied 1Hz to the right dlPFC, and 10Hz to the left dlPFC, at 100-120% MT. Weissman et al. (2018) found bilateral rTMS reduced STB in the active treatment arm, and this differed significantly to the sham arm (*p*=.020) and differed to their null finding for left dlPFC rTMS.“Treatments for the Prevention and Management of Suicide” (2019) found bilateral dlPFC rTMS reduced STB by 41%, which aligned closely with the 40.4% identified by Weissman et al. (2018) Remission was 25.8% for bilateral dlPFC rTMS (Weissman et al., 2018).

### dmPFC

One study investigated rTMS applied to the dmPFC, at a frequency of 5Hz (Calderón-Moctezuma et al., 2020). This study did not identify differences in STB after treatment across the active arm and sham arm, and did not report significant differences in STB within groups.

### Left, Right and Bilateral PFC

The retrospective study conducted by Abdelnaim et al. (2019) also investigated rTMS applied to the PFC more broadly. In addition to this, they report use of TMS to target the ACC, likely through dTMS. While their study indicated 47% showed STB improvement, 41.3% no STB change, 11.7% worsened STB, with one completed suicide during treatment, the results for each region were not independently reported, and interpretation of TMS efficacy at individual target sites was not possible.

### TMS to the prefrontal lobe (PFL)

#### Left PFL

One study investigated rTMS to the left PFL, at a frequency of 10Hz (Dai et al., 2020). They identified that left PFL rTMS had a significant reduction to STB at two (*p*<.050) and four weeks (*p*<.010). They also identified that the active arm had stronger reductions to STB than the sham arm, at both timepoints.

#### TMS to the left primary visual cortex

One study investigated iTBS applied to the left V1 at a frequency of 50Hz (Kong et al., 2023) STB decreased, with an 89% response rates, and a 71% remission rate. They found iTBS targeting the left V1 to be superior to that of the left dlPFC, although the difference was not significant.

#### Safety and Adverse Outcomes

Of the adverse events (AEs) reported, most were mild, transient and ultimately benign. Most common AEs included headaches (17 studies), dizziness (7 studies), discomfort or pain at the treatment site on the scalp (18 studies), or other mandibular and cephalic symptoms such as tooth pain (3 studies), jaw pain (2 studies), eye twitching/pain (2 studies), and changes to hearing (1 study) or vision (1 study). Three studies reported hypomanic episodes, although the studies were unclear on the pathogenesis (Bloch et al., 2008; Pan et al., 2023; Zhao et al., 2024). Notably, one participant experienced a syncopal event (Thai et al., 2024), and another experienced a seizure, the latter due to imprecise head-coil placement (Hickson et al., 2024). Finally, one study reported a case of second-degree burns as the result of improper TMS administration (George et al., 2014). Overall, the low number of significant AEs (.0009%) suggests that TMS is a low-risk treatment option when appropriate safety protocols are adhered to.

## Discussion and Conclusions

This review provides an updated summary investigating the use of various forms of TMS for the treatment of STB in adolescent and adult participants from 45 identified studies. Studies typically recruited individuals experiencing TRD and were treated for this indication, although studies investigating TMS treatment effects were also found for bipolar disorder, borderline personality disorder, PTSD, and TBI. Of the included studies, most were randomized controlled trials, with four retrospective studies and 16 open-label trials or open-label pilots. The left dlPFC was the most common site used for stimulation. In other studies, the left primary visual cortex and ACC were alternate target regions, as well as the PFC, bilateral dlPFC, and right dlPFC. Stimulation intensities were typically delivered at 100-120% of the resting motor threshold, although intensities as low at 80% were also reported (Bloch et al., 2008; M. H. Chen et al., 2022; Huang et al., 2025). Stimulation frequencies varied across studies, ranging from with low-frequency rTMS (1 Hz) to high-frequency rTMS up to 20 Hz. In addition, the number of pulses per session ranged from 400-8000, with heterogeneity in the number of rTMS sessions applied per day, the total number of sessions applied across a course of treatment. A range of outcome measures also used, depending on the conditions being treated. For the purpose of this review examining rTMS’s effects on treating STB, the majority of studies measured STB severity using the BSI, SSI and the C-SSRS rating scales.

Included studies generally reported significant reductions in STB, with most reporting between 47-97% response rates, particularly when receiving high frequency rTMS or iTBS to the left dlPFC. Worsening of symptoms was noted in 11.7-17% of participants with MDD in two studies (Abdelnaim et al., 2019; Wilkening et al., 2022), however, Abdelnaim et al. (2019) did not specify which TMS site or protocol(s) the increase of STB occurred in. The randomized controlled trials demonstrated the most consistent reductions in STB; while in many studies, both sham and active arms experienced decreases in STB, the active arms— particularly when administered to the left dlPFC—often showed superior results (Dai et al., 2022; Dai et al., 2020; Hines et al., 2022; Pan et al., 2018; Zhao et al., 2023). Interestingly, bilateral application of rTMS or iTBS to dlPFC also showed promise (Weissman et al., 2018), as did TMS applied to the visual cortex (Kong et al., 2023). In addition to the changes in STB, there were some differences noted in the efficacy of TMS when utilizing neuronavigation versus the “5cm rule”, with neuronavigated TMS providing more precise targeting of treatment locations, likely accounting for better outcomes (Sun et al., 2024).

It is important to note that some studies utilized TMS as an adjunct treatment to pharmacotherapy (Aaronson et al., 2024; Bozzay et al., 2020; Calderón-Moctezuma et al., 2020; Pan et al., 2018) or CBT (Adu et al., 2023). Other studies allowed for inclusions of individuals who had previous TMS (Abdelnaim et al., 2019) or alternate past treatments (Bozzay et al., 2020). While participants were often ineligible if they had previous ECT or TMS, Pan and colleagues (2018) found active rTMS combined with anti-depressant treatment was more effective than sham rTMS with anti-depressant treatment in reducing STB. Nonetheless, for those experiencing TRD (49% of included studies), findings supporting TMS’ efficacy and safety for STB are promising for long term management and treatment (Adu et al., 2023; Berlim et al., 2014; Croarkin & MacMaster, 2019; Croarkin et al., 2018; Wang et al., 2022; Wang et al., 2024; Zhang et al., 2021; Zhao et al., 2023). In addition, the number of participants included within each study was heterogenous, particularly when considering the inclusion of open-label pilots (*n*=3 to *n*=332). However, as this review aimed to update the evidence regarding the safety and efficacy of TMS, particularly across the various existing protocols, inclusion of these studies provided further context to the potential for TMS as a treatment option for those experiencing STB.

Regarding participant demographics, overall, there were more females included than males. This is in keeping with sex differences in prevalence of internalizing disorders such as depression (van Loo et al., 2023). Furthermore, most participants within this review had a mean age between 42-55. Studies have indicated that depressive problems may often follow a “U-shaped” pattern, whereby the are highest in adolescence/young adulthood, stagnate during mid-adulthood, and increase during older adulthood (Sutin et al., 2013).

### Strengths and Limitations

We present the most current review of TMS for STB, with a third of the studies published since the last identified systematic review (Aaronson et al., 2024; Adu et al., 2023; Hickson et al., 2024; Huang et al., 2025; Kong et al., 2023; Li et al., 2024; Pan et al., 2023; Sun et al., 2024; Terpstra et al., 2023; Thai et al., 2024; Wilkinson et al., 2023; Zhan et al., 2024; Zhao et al., 2024; Zhao et al., 2023).There are challenges in synthesizing studies with divergent TMS protocols and methodologies, and while we have endeavored to compare outcomes of like-for-like treatments to the best of our abilities, this may be a limitation of the broader TMS literature, and subsequently our review. While our objective was to investigate the potential for TMS to reduce STB, the inclusion of adolescents and adults within this review may be viewed as a limitation. As adolescents undergo significant neurodevelopment, this may augment (or potentially enhance) their receptibility to TMS treatment (Croarkin & MacMaster, 2019; Thai et al., 2024). As most adolescents who do not receive treatment for depression or STB during their youth often go on to experience lifelong internalizing problems and comorbid conditions, including youth in TMS investigation is critical to the prevention of long-term symptomatology (Colizzi et al., 2020; Thai et al., 2024). Further, most studies excluded individuals that did not indicate passive suicidality and therefore these findings may not be generalizable to individuals experiencing active suicidal ideation with intent.

### Implications and Future Directions

Overall, this review suggests that TMS is efficacious in the treatment of STB, within adults and adolescents, with low risk for adverse outcomes. Moreover, TMS appears effective in alleviating STB irrespective of diagnoses of mental and physical health conditions. Key to sound clinical care in populations presenting with STB, astute clinical risk assessment and management are paramount, to ensure patient welfare while consideration and provision of STB treatments are provisioned. In certain instances of high acuity or illness complexity, rTMS may not be an ideal firstline treatment for the primary presenting illness and the comorbid STB. Future research is needed to determine the efficacy of TMS treatment for STB independently.

Randomized controlled trials specifically designed to evaluate rTMS’ treatment effects on STB as a primary outcome are needed to validate this promising treatment approach. Future research can also investigate alternate TMS treatment sites for STB beyond the PFC, as brain regions implicated in maladaptive emotion regulation, impulsivity, negative urgency and emotional learning, such as the Cognitive Control Network (CCN) and Salience Network (SEN), are strongly implicated in STB (Bruno et al., 2023). In the meantime, the extant literature supports the notion that rTMS applied to treat TRD or another primary psychiatric disorder where STB is present, that reduction in STB can be achieved.

## Supplementary Materials

### Search and Screening Criteria

**Databases:** PubMed and Web of Science

**Search Terms:**

**PubMed:** (“transcranial magnetic stimulation”) AND (suicid*)

**Filters applied:** English, Humans.

29/01/25

137 results

**Web of Science:** Search all fields “transcranial magnetic stimulation” AND suicid* In English.

**Document type:** Article, i.e., not review article, meeting abstract, or book chapter 29/01/25

174 results

**Total:** 311

**Screening:**

1. Rayyan flagged 162 papers as duplicates (i.e., 81 duplicates) to be checked.
2. Auto-resolved based on exact match of DOI resolved 102 duplicates.
3. 60 resolved in Rayyan.
4. Manual duplicate screen identified with sort by title found one further duplicate where author had edited title to make link to pdf more publicly available.

**Total:** 229

## Data Availability

All data in this manuscript comes from published studies as it is a systematic review

## References

Aaronson, S. T., Goldwaser, E. L., Croarkin, P. E., Geske, J. R., LeMahieu, A., Sklar, J. H., & Kung, S. (2024). A Pilot Study of High-Frequency Transcranial Magnetic Stimulation for Bipolar Depression. J Clin Psychiatry, 85(2). 10.4088/JCP.23m15056

Abbar, M., Demattei, C., El-Hage, W., Llorca, P.-M., Samalin, L., Demaricourt, P., Gaillard, R., Courtet, P., Vaiva, G., Gorwood, P., Fabbro, P., & Jollant, F. (2022). Ketamine for the acute treatment of severe suicidal ideation: double blind, randomised placebo controlled trial. BMJ, 376, e067194. 10.1136/bmj-2021-067194

Abdelnaim, M. A., Langguth, B., Deppe, M., Mohonko, A., Kreuzer, P. M., Poeppl, T. B., Hebel, T., & Schecklmann, M. (2019). Anti-Suicidal Efficacy of Repetitive Transcranial Magnetic Stimulation in Depressive Patients: A Retrospective Analysis of a Large Sample. Front Psychiatry, 10, 929. 10.3389/fpsyt.2019.00929

Abdollahpour Ranjbar, H., Bakhshesh-Boroujeni, M., Farajpour-Niri, S., Hekmati, I., Habibi Asgarabad, M., & Eskin, M. (2024). An examination of the mediating role of maladaptive emotion regulation strategies in the complex relationship between interpersonal needs and suicidal behavior. Front Psychiatry, 15, 1301695. 10.3389/fpsyt.2024.1301695

Adu, M. K., Shalaby, R., Eboreime, E., Sapara, A., Lawal, M. A., Chew, C., Daubert, S., Urichuck, L., Surood, S., Li, D., Snaterse, M., Mach, M., Chue, P., Greenshaw, A. J., & Agyapong, V. I. O. (2023). Apparent Lack of Benefit of Combining Repetitive Transcranial Magnetic Stimulation with Internet-Delivered Cognitive Behavior Therapy for the Treatment of Resistant Depression: Patient-Centered Randomized Controlled Pilot Trial. Brain Sci, 13(2). 10.3390/brainsci13020293

An, H. M., Hsu, A., Ismail, R., & Bickart, K. (2025). Personalized TMS to the Frontal Pole Modulates Connectivity Within a Specific Amygdala Circuit in a Patient with Persistent Post-concussion Symptoms (P11-4.002). Neurology, 104(7_Supplement_1), 5119. doi:10.1212/WNL.0000000000212151

Argyelan, M., Lencz, T., Kang, S., Ali, S., Masi, P. J., Moyett, E., Joanlanne, A., Watson, P., Sanghani, S., Petrides, G., & Malhotra, A. K. (2021). ECT-induced cognitive side effects are associated with hippocampal enlargement. Translational Psychiatry, 11(1), 516. 10.1038/s41398-021-01641-y

Baeken, C., Duprat, R., Wu, G. R., De Raedt, R., & van Heeringen, K. (2017). Subgenual Anterior Cingulate-Medial Orbitofrontal Functional Connectivity in Medication-Resistant Major Depression: A Neurobiological Marker for Accelerated Intermittent Theta Burst Stimulation Treatment? Biol Psychiatry Cogn Neurosci Neuroimaging, 2(7), 556–565. 10.1016/j.bpsc.2017.01.001

Baeken, C., Wu, G. R., & van Heeringen, K. (2019). Placebo aiTBS attenuates suicidal ideation and frontopolar cortical perfusion in major depression. Transl Psychiatry, 9(1), 38. 10.1038/s41398-019-0377-x

Barredo, J., Bozzay, M. L., Primack, J. M., Schatten, H. T., Armey, M. F., Carpenter, L. L., & Philip, N. S. (2021). Translating Interventional Neuroscience to Suicide: It’s About Time. Biological Psychiatry, 89(11), 1073–1083. 10.1016/j.biopsych.2021.01.013

Berlim, M. T., Van den Eynde, F., Tovar-Perdomo, S., Chachamovich, E., Zangen, A., & Turecki, G. (2014). Augmenting antidepressants with deep transcranial magnetic stimulation (DTMS) in treatment-resistant major depression. World J Biol Psychiatry, 15(7), 570–578. 10.3109/15622975.2014.925141

Bloch, Y., Grisaru, N., Harel, E. V., Beitler, G., Faivel, N., Ratzoni, G., Stein, D., & Levkovitz, Y. (2008). Repetitive transcranial magnetic stimulation in the treatment of depression in adolescents: an open-label study. J ect, 24(2), 156–159. 10.1097/YCT.0b013e318156aa49

Bozzay, M. L., Primack, J., Barredo, J., & Philip, N. S. (2020). Transcranial magnetic stimulation to reduce suicidality - A review and naturalistic outcomes. J Psychiatr Res, 125, 106–112. 10.1016/j.jpsychires.2020.03.016

Braun, C., Bschor, T., Franklin, J., & Baethge, C. (2016). Suicides and Suicide Attempts during Long-Term Treatment with Antidepressants: A Meta-Analysis of 29 Placebo-Controlled Studies Including 6,934 Patients with Major Depressive Disorder. Psychother Psychosom, 85(3), 171–179. 10.1159/000442293

Bruno, S., Anconetani, G., Rogier, G., Del Casale, A., Pompili, M., & Velotti, P. (2023). Impulsivity traits and suicide related outcomes: A systematic review and meta-analysis using the UPPS model. J Affect Disord, 339, 571–583. 10.1016/j.jad.2023.07.086

Bruton, A. M., Wesemann, D. G., Machingo, T. A., Majak, G., Johnstone, J. M., & Marshall, R. D. (2025). Ketamine for mood disorders, anxiety, and suicidality in children and adolescents: a systematic review. European Child & Adolescent Psychiatry, 34(1), 141–157. 10.1007/s00787-024-02458-y

Calderón-Moctezuma, A. R., Reyes-López, J. V., Rodríguez-Valdés, R., Barbosa-Luna, M., Ricardo-Garcell, J., Espino-Cortés, M., Hernández-Chan, N., García-Noguez, L., Roque-Roque, G., Trejo-Cruz, G., Cañizares-Gómez, S., & Hernández-Montiel, H. (2020). Improvement in borderline personality disorder symptomatology after repetitive transcranial magnetic stimulation of the dorsomedial prefrontal cortex: preliminary results. Braz J Psychiatry, 43(1), 65–69. 10.1590/1516-4446-2019-0591

Caudle, M. M., Dugas, N. N., Patel, K., Moore, R. C., Thomas, M. L., & Bomyea, J. (2024). Repetitive negative thinking as a unique transdiagnostic risk factor for suicidal ideation. Psychiatry Research, 334, 115787. 10.1016/j.psychres.2024.115787

Chen, G.-W., Hsu, T.-W., Ching, P.-Y., Pan, C.-C., Chou, P.-H., & Chu, C.-S. (2022). Efficacy and Tolerability of Repetitive Transcranial Magnetic Stimulation on Suicidal Ideation: A Systemic Review and Meta-Analysis [Systematic Review]. Frontiers in Psychiatry, 13. 10.3389/fpsyt.2022.884390

Chen, M. H., Cheng, C. M., Li, C. T., Tsai, S. J., Lin, W. C., Bai, Y. M., & Su, T. P. (2022). Comparative study of low-dose ketamine infusion and repetitive transcranial magnetic stimulation in treatment-resistant depression: A posthoc pooled analysis of two randomized, double-blind, placebo-controlled studies. Psychiatry Res, 316, 114749. 10.1016/j.psychres.2022.114749

Civardi, S. C., Besana, F., Carnevale Miacca, G., Mazzoni, F., Arienti, V., Politi, P., Brondino, N., & Olivola, M. (2024). Risk factors for suicidal attempts in a sample of outpatients with treatment-resistant depression: an observational study [Original Research]. Frontiers in Psychiatry, Volume 15 - 2024. 10.3389/fpsyt.2024.1371139

Cohen, S. L., Bikson, M., Badran, B. W., & George, M. S. (2022). A visual and narrative timeline of US FDA milestones for Transcranial Magnetic Stimulation (TMS) devices. Brain Stimul, 15(1), 73–75. 10.1016/j.brs.2021.11.010

Colizzi, M., Lasalvia, A., & Ruggeri, M. (2020). Prevention and early intervention in youth mental health: is it time for a multidisciplinary and trans-diagnostic model for care? International Journal of Mental Health Systems, 14(1), 23. 10.1186/s13033-020-00356-9

Croarkin, P. E., & MacMaster, F. P. (2019). Transcranial Magnetic Stimulation for Adolescent Depression. Child Adolesc Psychiatr Clin N Am, 28(1), 33–43. 10.1016/j.chc.2018.07.003

Croarkin, P. E., Nakonezny, P. A., Deng, Z. D., Romanowicz, M., Voort, J. L. V., Camsari, D. D., Schak, K. M., Port, J. D., & Lewis, C. P. (2018). High-frequency repetitive TMS for suicidal ideation in adolescents with depression. J Affect Disord, 239, 282–290. 10.1016/j.jad.2018.06.048

Dai, L., Wang, P., Du, H., Guo, Q., Li, F., He, X., & Zou, S. (2022). High-frequency Repetitive Transcranial Magnetic Stimulation (rTMS) Accelerates onset Time of Beneficial Treating Effects and Improves Clinical Symptoms of Depression. CNS Neurol Disord Drug Targets, 21(6), 500–510. 10.2174/1871527320666211104123343

Dai, L., Wang, P., Zhang, P., Guo, Q., Du, H., Li, F., He, X., & Luan, R. (2020). The therapeutic effect of repetitive transcranial magnetic stimulation in elderly depression patients. Medicine (Baltimore), 99(32), e21493. 10.1097/md.0000000000021493

Davila, M. C., Ely, B., & Manzardo, A. M. (2019). Repetitive transcranial magnetic stimulation (rTMS) using different TMS instruments for major depressive disorder at a suburban tertiary clinic. Ment Illn, 11(1), 7947. 10.4081/mi.2019.7947

Del Matto, L., Muscas, M., Murru, A., Verdolini, N., Anmella, G., Fico, G., Corponi, F., Carvalho, A. F., Samalin, L., Carpiniello, B., Fagiolini, A., Vieta, E., & Pacchiarotti, I. (2020). Lithium and suicide prevention in mood disorders and in the general population: A systematic review. Neuroscience & Biobehavioral Reviews, 116, 142–153. 10.1016/j.neubiorev.2020.06.017

Desmyter, S., Duprat, R., Baeken, C., Van Autreve, S., Audenaert, K., & van Heeringen, K. (2016). Accelerated Intermittent Theta Burst Stimulation for Suicide Risk in Therapy-Resistant Depressed Patients: A Randomized, Sham-Controlled Trial. Front Hum Neurosci, 10, 480. 10.3389/fnhum.2016.00480

Fehling, K. B., & Selby, E. A. (2020). Suicide in DSM-5: Current Evidence for the Proposed Suicide Behavior Disorder and Other Possible Improvements. Front Psychiatry, 11, 499980. 10.3389/fpsyt.2020.499980

Firth, J., Solmi, M., Wootton, R. E., Vancampfort, D., Schuch, F. B., Hoare, E., Gilbody, S., Torous, J., Teasdale, S. B., Jackson, S. E., Smith, L., Eaton, M., Jacka, F. N., Veronese, N., Marx, W., Ashdown-Franks, G., Siskind, D., Sarris, J., Rosenbaum, S.,… Stubbs, B. (2020). A meta-review of “lifestyle psychiatry”: the role of exercise, smoking, diet and sleep in the prevention and treatment of mental disorders. World Psychiatry, 19(3), 360–380. 10.1002/wps.20773

George, M. S., Raman, R., Benedek, D. M., Pelic, C. G., Grammer, G. G., Stokes, K. T., Schmidt, M., Spiegel, C., Dealmeida, N., Beaver, K. L., Borckardt, J. J., Sun, X., Jain, S.,& Stein, M. B. (2014). A two-site pilot randomized 3 day trial of high dose left prefrontal repetitive transcranial magnetic stimulation (rTMS) for suicidal inpatients. Brain Stimul, 7(3), 421–431. 10.1016/j.brs.2014.03.006

Hadley, D., Anderson, B. S., Borckardt, J. J., Arana, A., Li, X., Nahas, Z.,& George, M. S. (2011). Safety, tolerability, and effectiveness of high doses of adjunctive daily left prefrontal repetitive transcranial magnetic stimulation for treatment-resistant depression in a clinical setting. J ect, 27(1), 18–25. 10.1097/YCT.0b013e3181ce1a8c

Han, S., Li, X. X., Wei, S., Zhao, D., Ding, J., Xu, Y., Yu, C., Chen, Z., Zhou, D. S.,& Yuan, T. F. (2023). Orbitofrontal cortex-hippocampus potentiation mediates relief for depression: A randomized double-blind trial and TMS-EEG study. Cell Rep Med, 4(6), 101060. 10.1016/j.xcrm.2023.101060

Hickson, R., Simonsen, M. W., Miller, K. J.,& Madore, M. R. (2024). Durability of deep transcranial magnetic stimulation for veterans with treatment resistant depression with comorbid suicide risk and PTSD symptoms. Psychiatry Res, 332, 115690. 10.1016/j.psychres.2023.115690

Hines, C. E., Mooney, S., Watson, N. L., Looney, S. W.,& Wilkie, D. J. (2022). Repetitive Transcranial Magnetic Stimulation Promotes Rapid Psychiatric Stabilization in Acutely Suicidal Military Service Members. J ect, 38(2), 103–109. 10.1097/yct.0000000000000810

Huang, X., Xi, C., Fang, Y., Ye, R., Wang, X., Zhang, S., Cui, Y., Guo, Y., Zhang, J., Ji, G.-J., Zhu, C., Luo, Y., Chen, X., Wang, K., Tian, Y.,& Yu, F. (2025). Therapeutic Efficacy of Reward Circuit-Targeted Transcranial Magnetic Stimulation (TMS) on Suicidal Ideation in Depressed Patients: A Sham-Controlled Trial of Two TMS Protocols. Depression and Anxiety, 2025(1), 1767477. 10.1155/da/1767477

Iannucci, J.,& Nierenberg, B. (2022). Suicide and suicidality in children and adolescents with chronic illness: A systematic review. Aggression and Violent Behavior, 64, 101581. 10.1016/j.avb.2021.101581

Ingrosso, G., Cleare, A. J.,& Juruena, M. F. (2025). Is there a risk of addiction to ketamine during the treatment of depression? A systematic review of available literature. Journal of Psychopharmacology, 39(1), 49–65. 10.1177/02698811241303597

Jørgensen, A., Gronemann, F. H., Rozing, M. P., Jørgensen, M. B.,& Osler, M. (2024). Clinical Outcomes of Continuation and Maintenance Electroconvulsive Therapy. JAMA Psychiatry, 81(12), 1207–1214. 10.1001/jamapsychiatry.2024.2360

Keshtkar, M., Ghanizadeh, A.,& Firoozabadi, A. (2011). Repetitive transcranial magnetic stimulation versus electroconvulsive therapy for the treatment of major depressive disorder, a randomized controlled clinical trial. J ect, 27(4), 310–314. 10.1097/YCT.0b013e318221b31c

Kong, Y., Zhou, J., Zhao, M., Zhang, Y., Tan, T., Xu, Z., Hou, Z., Yuan, Y., Tan, L., Song, R., Shi, Y., Feng, H., Wu, W., Zhao, Y.,& Zhang, Z. (2023). Non-inferiority of intermittent theta burst stimulation over the left V(1) vs. classical target for depression: A randomized, double-blind trial. J Affect Disord, 343, 59–70. 10.1016/j.jad.2023.09.024

Lascelles, K., Marzano, L., Brand, F., Trueman, H., McShane, R.,& Hawton, K. (2021). Ketamine treatment for individuals with treatment-resistant depression: longitudinal qualitative interview study of patient experiences. BJPsych Open, 7(1), e9.

Li, B., Zhao, N., Tang, N., Friston, K. J., Zhai, W., Wu, D., Liu, J., Chen, Y., Min, Y., Qiao, Y., Liu, W., Shu, W., Liu, M., Zhou, P., Guo, L., Qi, S., Cui, L.-B., & Wang, H. (2024). Targeting suicidal ideation in major depressive disorder with MRI-navigated Stanford accelerated intelligent neuromodulation therapy. Translational Psychiatry, 14(1), 21. 10.1038/s41398-023-02707-9

Lusicic, A., Schruers, K. R., Pallanti, S., & Castle, D. J. (2018). Transcranial magnetic stimulation in the treatment of obsessive-compulsive disorder: current perspectives. Neuropsychiatr Dis Treat, 14, 1721–1736. 10.2147/ndt.S121140

Mehta, S., Downar, J., Mulsant, B. H., Voineskos, D., Daskalakis, Z. J., Weissman, C. R., Vila-Rodriguez, F., & Blumberger, D. M. (2022). Effect of high frequency versus theta-burst repetitive transcranial magnetic stimulation on suicidality in patients with treatment-resistant depression. Acta Psychiatr Scand, 145(5), 529–538. 10.1111/acps.13412

Mutz, J., Edgcumbe, D. R., Brunoni, A. R., & Fu, C. H. Y. (2018). Efficacy and acceptability of non-invasive brain stimulation for the treatment of adult unipolar and bipolar depression: A systematic review and meta-analysis of randomised sham-controlled trials. Neuroscience & Biobehavioral Reviews, 92, 291–303. 10.1016/j.neubiorev.2018.05.015

Ozcan, S., Gica, S., & Gulec, H. (2020). Suicidal behavior in treatment resistant major depressive disorder patients treated with transmagnetic stimulation(TMS) and its relationship with cognitive functions. Psychiatry Res, 286, 112873. 10.1016/j.psychres.2020.112873

Page, M. J., McKenzie, J. E., Bossuyt, P. M., Boutron, I., Hoffmann, T. C., Mulrow, C. D., Shamseer, L., Tetzlaff, J. M., Akl, E. A., Brennan, S. E., Chou, R., Glanville, J., Grimshaw, J. M., Hróbjartsson, A., Lalu, M. M., Li, T., Loder, E. W., Mayo-Wilson, E., McDonald, S.,… Moher, D. (2021). The PRISMA 2020 statement: an updated guideline for reporting systematic reviews. BMJ, 372, n71. 10.1136/bmj.n71

Page, M. J., Moher, D., Bossuyt, P. M., Boutron, I., Hoffmann, T. C., Mulrow, C. D., Shamseer, L., Tetzlaff, J. M., Akl, E. A., Brennan, S. E., Chou, R., Glanville, J., Grimshaw, J. M., Hróbjartsson, A., Lalu, M. M., Li, T., Loder, E. W., Mayo-Wilson, E., McDonald, S.,… McKenzie, J. E. (2021). PRISMA 2020 explanation and elaboration: updated guidance and exemplars for reporting systematic reviews. BMJ, 372, n160. 10.1136/bmj.n160

Pan, F., Li, D., Wang, X., Lu, S., Xu, Y., & Huang, M. (2018). Neuronavigation-guided high-dose repetitive transcranial magnetic stimulation for the treatment of depressive adolescents with suicidal ideation: a case series. Neuropsychiatr Dis Treat, 14, 2675–2679. 10.2147/ndt.S176125

Pan, F., Mou, T., Shao, J., Chen, H., Tao, S., Wang, L., Jiang, C., Zhao, M., Wang, Z., Hu, S., Xu, Y., & Huang, M. (2023). Effects of neuronavigation-guided rTMS on serum BDNF, TrkB and VGF levels in depressive patients with suicidal ideation. J Affect Disord, 323, 617–623. 10.1016/j.jad.2022.11.059

Petrosino, N. J., Wout-Frank, M. V., Aiken, E., Swearingen, H. R., Barredo, J., Zandvakili, A., & Philip, N. S. (2020). One-year clinical outcomes following theta burst stimulation for post-traumatic stress disorder. Neuropsychopharmacology, 45(6), 940–946. 10.1038/s41386-019-0584-4

Porter, R. J., Baune, B. T., Morris, G., Hamilton, A., Bassett, D., Boyce, P., Hopwood, M. J., Mulder, R., Parker, G., Singh, A. B., Outhred, T., Das, P., & Malhi, G. S. (2020). Cognitive side-effects of electroconvulsive therapy: what are they, how to monitor them and what to tell patients. BJPsych Open, 6(3), e40. 10.1192/bjo.2020.17

Rao, V., Bechtold, K., McCann, U., Roy, D., Peters, M., Vaishnavi, S., Yousem, D., Mori, S., Yan, H., Leoutsakos, J., Tibbs, M., & Reti, I. (2019). Low-Frequency Right Repetitive Transcranial Magnetic Stimulation for the Treatment of Depression After Traumatic Brain Injury: A Randomized Sham-Controlled Pilot Study. J Neuropsychiatry Clin Neurosci, 31(4), 306–318. 10.1176/appi.neuropsych.17110338

Salik, I., & Marwaha, R. (2025). Electroconvulsive Therapy. In StatPearls. StatPearls Publishing Copyright © 2025, StatPearls Publishing LLC.

Shahsavar, Y., & Choudhury, A. (2025). Effectiveness of evidence based mental health apps on user health outcome: A systematic literature review. PLoS One, 20(3), e0319983. 10.1371/journal.pone.0319983

Sienaert, P., Brus, O., Lambrichts, S., Lundberg, J., Nordanskog, P., Obbels, J., Verspecht, S., Vansteelandt, K., & Nordenskjöld, A. (2022). Suicidal ideation and ECT, ECT and suicidal ideation: A register study. Acta Psychiatr Scand, 146(1), 74–84. 10.1111/acps.13425

Soleimani, G., Conelea, C. A., Kuplicki, R., Opitz, A., Lim, K. O., Paulus, M. P., & Ekhtiari, H. (2025). Targeting VMPFC-amygdala circuit with TMS in substance use disorder: A mechanistic framework. Addict Biol, 30(1), e70011. 10.1111/adb.70011

Sun, Y., Liu, X., Li, Y., Zhi, Q., & Xia, Y. (2024). Effectiveness of individualized rTMS under sMRI guidance in reducing depressive symptoms and suicidal ideation in adolescents with depressive disorders: an open-label study. Front Psychiatry, 15, 1485878. 10.3389/fpsyt.2024.1485878

Sutin, A. R., Terracciano, A., Milaneschi, Y., An, Y., Ferrucci, L., & Zonderman, A. B. (2013). The trajectory of depressive symptoms across the adult life span. JAMA Psychiatry, 70(8), 803–811. 10.1001/jamapsychiatry.2013.193

Tang, N., Sun, C., Wang, Y., Li, X., Liu, J., Chen, Y., Sun, L., Rao, Y., Li, S., Qi, S., & Wang, H. (2021). Clinical Response of Major Depressive Disorder Patients With Suicidal Ideation to Individual Target-Transcranial Magnetic Stimulation. Front Psychiatry, 12, 768819. 10.3389/fpsyt.2021.768819

Terpstra, A. R., Vila-Rodriguez, F., LeMoult, J., Chakrabarty, T., Nair, M., Humaira, A., Gregory, E. C., & Todd, R. M. (2023). Cognitive-affective processes and suicidality in response to repetitive transcranial magnetic stimulation for treatment resistant depression. J Affect Disord, 321, 182–190. 10.1016/j.jad.2022.10.041

Thai, M., Nair, A. U., Klimes-Dougan, B., Albott, C. S., Silamongkol, T., Corkrum, M., Hill, D., Roemer, J. W., Lewis, C. P., Croarkin, P. E., Lim, K. O., Widge, A. S., Nahas, Z., Eberly, L. E., & Cullen, K. R. (2024). Deep transcranial magnetic stimulation for adolescents with treatment-resistant depression: A preliminary dose-finding study exploring safety and clinical effectiveness. Journal of Affective Disorders, 354, 589–600. 10.1016/j.jad.2024.03.061

Treatments for the Prevention and Management of Suicide. (2019). Annals of Internal Medicine, 171(5), 334–342. 10.7326/m19-0869%m31450239

van Loo, H. M., Beijers, L., Wieling, M., de Jong, T. R., Schoevers, R. A., & Kendler, K. S. (2023). Prevalence of internalizing disorders, symptoms, and traits across age using advanced nonlinear models. Psychol Med, 53(1), 78–87. 10.1017/s0033291721001148

Wall, C. A., Croarkin, P. E., Sim, L. A., Husain, M. M., Janicak, P. G., Kozel, F. A., Emslie, G. J., Dowd, S. M., & Sampson, S. M. (2011). Adjunctive use of repetitive transcranial magnetic stimulation in depressed adolescents: a prospective, open pilot study. J Clin Psychiatry, 72(9), 1263–1269. 10.4088/JCP.11m07003

Wang, Q., Huang, H., Li, D., Wang, Y., Qi, N., Ci, Y., & Xu, T. (2022). Intensive rTMS for treatment-resistant depression patients with suicidal ideation: An open-label study. Asian J Psychiatr, 74, 103189. 10.1016/j.ajp.2022.103189

Wang, Q., Li, L., Zhao, H., Cheng, W., Cui, G., Fan, L., Dong, X., Xu, T., & Geng, Z. (2024). Predictors of response to accelerated rTMS in the treatment of treatment-resistant depression. Eur Arch Psychiatry Clin Neurosci. 10.1007/s00406-024-01903-y

Weissman, C. R., Blumberger, D. M., Brown, P. E., Isserles, M., Rajji, T. K., Downar, J., Mulsant, B. H., Fitzgerald, P. B., & Daskalakis, Z. J. (2018). Bilateral Repetitive Transcranial Magnetic Stimulation Decreases Suicidal Ideation in Depression. J Clin Psychiatry, 79(3). 10.4088/JCP.17m11692

Wilkening, J., Witteler, F., & Goya-Maldonado, R. (2022). Suicidality and relief of depressive symptoms with intermittent theta burst stimulation in a sham-controlled randomized clinical trial. Acta Psychiatr Scand, 146(6), 540–556. 10.1111/acps.13502

Wilkinson, S. T., Ballard, E. D., Bloch, M. H., Mathew, S. J., Murrough, J. W., Feder, A., Sos, P., Wang, G., Zarate, C. A., Jr., & Sanacora, G. (2018). The Effect of a Single Dose of Intravenous Ketamine on Suicidal Ideation: A Systematic Review and Individual Participant Data Meta-Analysis. Am J Psychiatry, 175(2), 150–158. 10.1176/appi.ajp.2017.17040472

Wilkinson, S. T., Diaz, D. T., Rupp, Z. W., Kidambi, A., Ramirez, K. L., Flores, J. M., Avila-Quintero, V. J., Rhee, T. G., Olfson, M., & Bloch, M. H. (2023). Pharmacological and Somatic Treatment Effects on Suicide in Adults: A Systematic Review and Meta-Analysis. Focus, 21(2), 197–208. 10.1176/appi.focus.23021006

*World Health Organization*. (2024). Retrieved March 14 2025 from https://www.who.int/news-room/fact-sheets/detail/suicide

Yesavage, J. A., Fairchild, J. K., Mi, Z., Biswas, K., Davis-Karim, A., Phibbs, C. S., Forman, S. D., Thase, M., Williams, L. M., Etkin, A., O’Hara, R., Georgette, G., Beale, T., Huang, G. D., Noda, A., & George, M. S. (2018). Effect of Repetitive Transcranial Magnetic Stimulation on Treatment-Resistant Major Depression in US Veterans: A Randomized Clinical Trial. JAMA Psychiatry, 75(9), 884–893. 10.1001/jamapsychiatry.2018.1483

Zapf, L., Kaster, T. S., Vila-Rodriguez, F., Daskalakis, Z. J., Downar, J., & Blumberger, D. M. (2024). The effect of once-daily vs. twice-daily intermittent theta burst stimulation on suicidal ideation in treatment-resistant depression. Eur Arch Psychiatry Clin Neurosci. 10.1007/s00406-024-01929-2

Zhan, D., Gregory, E. C., Humaira, A., Wong, H., Klonsky, E. D., Levit, A., Ridgway, L., & Vila-Rodriguez, F. (2024). Trajectories of suicidal ideation during rTMS for treatment-resistant depression. J Affect Disord, 360, 108–113. 10.1016/j.jad.2024.05.109

Zhang, T., Zhu, J., Wang, J., Tang, Y., Xu, L., Tang, X., Hu, Y., Wei, Y., Cui, H., Liu, X., Hui, L., Li, C., & Wang, J. (2021). An Open-label Trial of Adjuvant High-frequency Left Prefrontal Repetitive Transcranial Magnetic Stimulation for Treating Suicidal Ideation in Adolescents and Adults With Depression. J ect, 37(2), 140–146. 10.1097/yct.0000000000000739

Zhao, H., Jiang, C., Zhao, M., Ye, Y., Yu, L., Li, Y., Luan, H., Zhang, S., Xu, P., Chen, X., Pan, F., Shang, D., Hu, X., Jin, K., Chen, J., Mou, T., Hu, S., Fitzgibbon, B. M., Fitzgerald, P. B.,… Huang, M. (2024). Comparisons of Accelerated Continuous and Intermittent Theta Burst Stimulation for Treatment-Resistant Depression and Suicidal Ideation. Biol Psychiatry, 96(1), 26–33. 10.1016/j.biopsych.2023.12.013

Zhao, Y., He, Z., Luo, W., Yu, Y., Chen, J., Cai, X., Gao, J., Li, L., Gao, Q., Chen, H., & Lu, F. (2023). Effect of intermittent theta burst stimulation on suicidal ideation and depressive symptoms in adolescent depression with suicide attempt: A randomized sham-controlled study. J Affect Disord, 325, 618–626. 10.1016/j.jad.2023.01.061

Zisook, S., Domingues, I., & Compton, J. (2023). Pharmacologic Approaches to Suicide Prevention. Focus (Am Psychiatr Publ), 21(2), 137–144. 10.1176/appi.focus.20220076

